# A Pilot Study of Stereoelectroencephalography Electrodes in a Patient with Refractory Chronic Migraine: Personalized Targets for Precise Deep Brain Stimulation

**DOI:** 10.1101/2023.06.19.23291563

**Authors:** Hulin Zhao, Shuhua Zhang, Yining Wang, Chuting Zhang, Zihua Gong, Mingjie Zhang, Wei Dai, Ye Ran, Wenbin Shi, Yuanyuan Dang, Aijun Liu, Zhengbo Zhang, Chien-Hung Yeh, Zhao Dong

## Abstract

**Background and Objectives:** The complexity of neural circuits and the heterogeneity of brain networks are barriers for further improving the efficacy of DBS. This study aimed to establish a clinical paradigm to personalize the design of DBS in patients with refractory headache, which would constitute a milestone in this field.

**Methods:** We implanted 14 stereoelectroencephalography electrodes in a patient with refractory migraine for clinical monitoring and electrophysiological recording. During monitoring, we collected the VAS score in 5-min increments, and recorded electrophysiological data in real-time. Data were classified into two types of symptoms (high and low symptoms) for determining the spectral power features of specific brain regions reflecting pain fluctuations, which we called Biomarker, using statistical analyses and cross-validated machine-learning models. During stimulation, we tested the clinical effect through a systematic bipolar stimulation survey and blinded sham-controlled stimulation studies, and collected real-time electrophysiological data. Based on the identification of brain areas with clinical improvement, the optimal target for stimulate was determined by validating the clinical response against the biomarker, and phase-amplitude coupling finally.

**Results:** For biomarker, RNAc-HFO was the most considerably correlated with VAS score (rho = 0.5292, *p* < 0.0001), and differed significantly between mild and severe pain levels (*p* = 0.0003), also with the greatest weighting in the characteristic ranking. The machine-learning model showed an accuracy and AUC remaining at 75.51% and 0.77, respectively, for RAC-HFO. For target, LdACC was identified as the most effective stimulation target, based on the VAS score reported over the stimulation period. VAS score (*p* = 0.006), RNAc-HFO (*p* = 0.0029) were significantly improved after stimulation compared to pre-stimulation in LdACC. The significant modulatory effect of RNAc-HFO by the low-frequency phase of LdACC also confirmed the modulatory effect of LdACC and RNAc during headache fluctuation.

**Discussion:** As a pilot study for exploring precise and personalized DBS in refractory migraine, we identified the biomarker and optimal target via the integration of clinical and electrophysiological data. The concept of the proposed data-driven approach to optimizing personalized treatment strategies for DBS may create a new frontier in the field of refractory headache and pain.

## Introduction

Migraine is a common neurovascular disorder and the second leading cause of disability worldwide according to the 2016 Global Burden of Disease ^1, 2^. Chronic migraine, defined as a headache occurring on ≥ 15 days per month for > 3 months with migraine occurring on ≥ 8 of these days ^3^, is associated with even greater disability, indicating the need for effective treatment. The current conventional treatments include both pharmacological and non-pharmacological therapies, but approximately 5.1% of patients with refractory chronic migraine (rCM) showed minimal or no response to standard and/or active treatment ^4^. Therefore, given the low quality of life, huge socioeconomic burden, and high treatment needs of patients with rCM, investigating more effective treatment regimens is critical.

Deep brain stimulation (DBS) is a well-established therapy for treatment-resistant conditions such as movement disorders, psychiatric disorders, epilepsy, and pain syndromes (e.g., neuropathic pain and cluster headache) ^5^. A recent meta-analysis showed that DBS for treatment of refractory chronic cluster headache (rCCH) had a pooled response rate of only 77%, with high heterogeneity of results ^6^. Moreover, the only randomized placebo-controlled double-blind trial of DBS in patients with rCCH to date did not support the efficacy of this treatment ^7^. Previous studies have shown that the variable efficacy of DBS for rCCH may be due not only to misalignment of electrodes but also to the non-individualized stimulation target, the complicated pathophysiological or disease state, and the individual structure or functional anatomy of the patient ^6, 8^. In addition to these issues, the multi-oscillatory neural dynamics and the heterogeneity of neural circuits in the brain have made optimizing and allocating individualized stimulation regimens for precise treatment of headaches by DBS a major challenge. Some recent studies of DBS for patients with treatment-resistant depression have explored individualized targets with stereoelectroencephalography (SEEG) electrodes ^9, 10^, providing encouraging findings regarding precise treatment of refractory headaches with DBS. To the best of our knowledge, however, no case reports have described the precise treatment of refractory headaches or even pain by integrating clinical assessment and electrophysiological data. Therefore, we hypothesized that SEEG recordings can be used to access headache intensity-linked biomarkers that will aid in brain region identification, identifying specific targets for the first application of DBS for rCM to date **(Figure 1)**.

**Figure 1.**
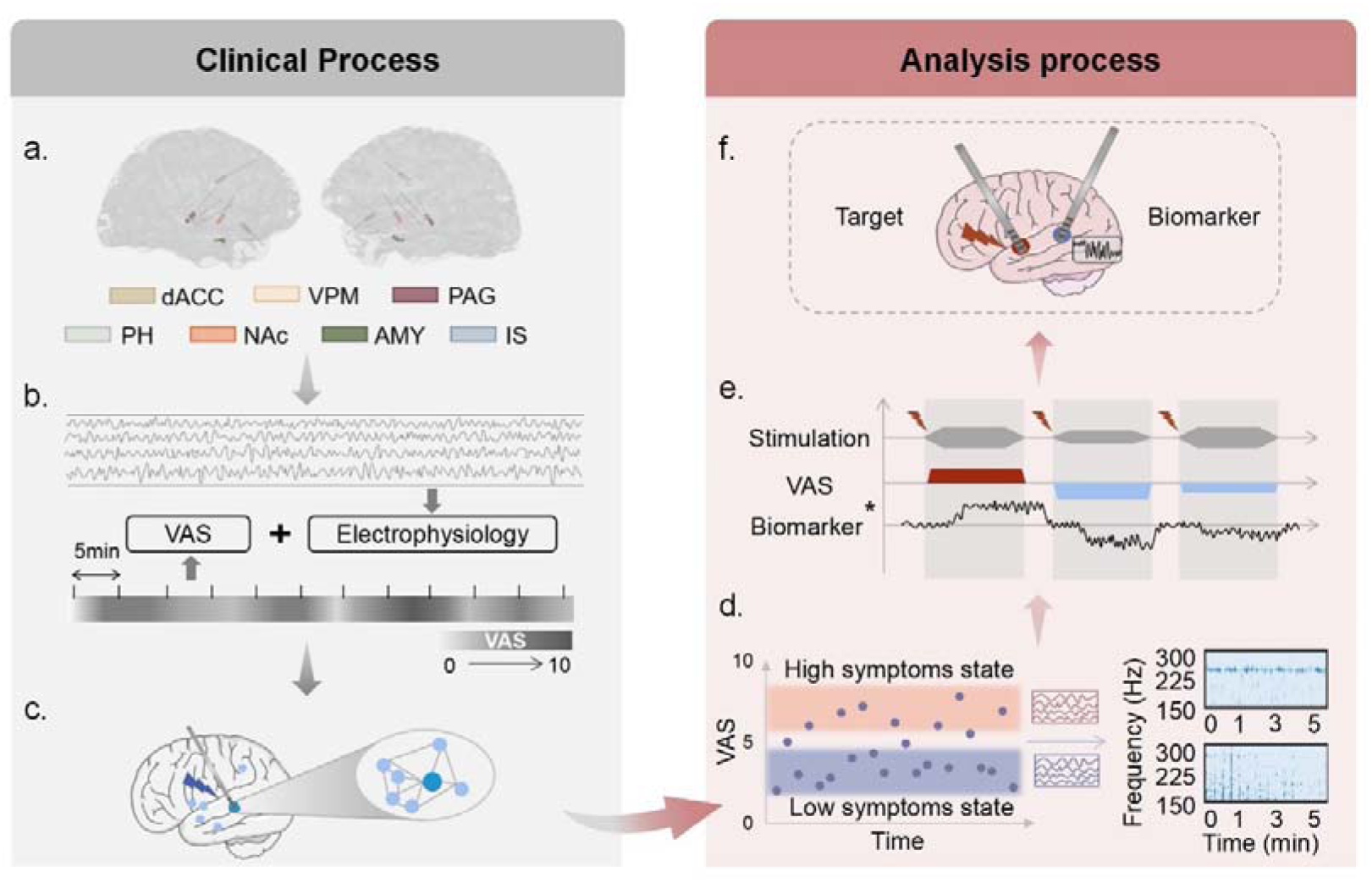
Study procedure. (a) Implantation and visualization of the 14 SEEG electrodes. **(b)** Clinical monitoring and electrophysiological recordings. Headache severity was recorded in 5-minute increments using a visual analog scale (VAS) and real-time electrophysiological recordings (SEEG). The VAS was used to rate the intensity of the headache and ranged from 0 to 10, with 0 representing no pain and 10 representing the most severe pain. **(c)** Stimulations with different parameters were applied to the effective contacts of all brain regions for clinical effects, thereby altering the functional connectivity among brain regions (dark blue represents the stimulated brain region, and light blue represents the brain regions with a pronounced change in response to the headache intensity). The SEEG data, VAS scores, and other responses (e.g., physiological responses induced by the stimulation or headache-related concomitant symptoms) were all recorded simultaneously in real time during stimulation. **(d)** Clinical and electrophysiological features from the monitoring period were combined to determine the biomarker responding to the headache fluctuation. According to the VAS assessment and SEEG data, the intensity of headache was divided into two states (pink: high-symptom state; blue: low-symptom state), and biomarkers determined by real-time SEEG analyses reflected the various headache states. **(e)** Clinical and electrophysiological data from the monitoring period were combined to determine the best stimulating target for maximum headache relief. For the stimulus data, fluctuations in headache intensity during stimulation (exacerbation, no change, or improvement) were matched with corresponding changes in the biomarker to determine the best target, supported by both the clinical response and electrophysiology. Red indicates a worsening headache, whereas blue indicates the opposite. The line width in the stimulation denotes the stimulus intensity. *Biomarker refers to the spectral power feature that optimally reflects varying headache states. **(f)** The optimization of both the stimulation targets and biomarkers served to initiate biomarker-directed personalized stimulation of DBS. SEEG: stereoelectroencephalography; VAS: visual analog scale; PAG/PVG: periaqueductal/periventricular gray area; VPM/VPL: ventral posteromedial/ventral posterolateral; NAc: nucleus accumbens; dACC: dorsal anterior cingulate cortex; PH: posterior hypothalamus; AMY: amygdala; IS: insula

## Material and methods

### Participant

The patient was a right-handed man with a more than 5 years history of bilateral temporal throbbing headaches. The migraine attacks were assigned a VAS score of 2 to 6, an attack frequency of two to three times per week, and a duration of 4 to 12 hours. The attacks were often accompanied by nausea, vomiting, photophobia, phonophobia, and a variety of autonomic symptoms (e.g., moist eyes, tearing, and yawning), and they disrupted the patient’s daily activities. The number of attacks had gradually increased until 3 years before presentation, at which time the patient developed a constant headache with a VAS score of 3 to 4 and several exacerbations per day, almost always manifesting as migraines. Furthermore, the exacerbations were always accompanied by typical triggers. He sought treatment at several hospitals and was eventually diagnosed with chronic migraine. He began a long course of both acute and preventive care, including two hospitalizations at our facility. During his initial hospitalization, he underwent necessary tests such as cranial magnetic resonance imaging and lumbar puncture, both of which produced essentially normal results and ruled out secondary headaches. The psychology department simultaneously assisted in excluding the possibility of anxiety or depression. We spent 3 months adjusting his preventive treatment regimen, but his symptoms only slightly improved. During his second hospitalization, given the long disease course and treatment resistance, we further adjusted his regimen and tried other new medications available to him; however, he gained no significant relief from his symptoms. Overall, the patient received all available preventive treatments (antidepressants, antiepileptics, beta-blockers, calcium channel blockers, and onabotulinumtoxin A) at appropriate doses and durations, as well as adequate non-pharmacological treatments, with no improvement; therefore, we considered his condition to be consistent with refractory migraine ^11^. Following a thorough evaluation and observation, the patient underwent SEEG in preparation for precise DBS treatment.

### Surgical procedure

The patient underwent the SEEG electrode implantation procedure as follows. According to previous research, we implanted 14 SEEG electrodes (Beijing Sinovation Medical Technology Co., Ltd., Beijing, China) in the bilateral periaqueductal/periventricular gray area (PAG/PVG), ventral posteromedial and posterolateral (VPM/VPL), NAc, dACC, posterior hypothalamus, amygdala (AMY), and insula (IS), which have shown the most promise for symptomatic improvement ^12, 13^ (**Figure 1a**). The surgical plan was created with Reme-Studio, and the procedure was carried out with the assistance of a Remebot neurosurgical robot (Beijing Baihui Weikang Technology Co., Ltd., Beijing, China). The electrode positions were confirmed intraoperatively using computed tomography. No complications occurred during the procedure. The electrodes were completely removed after 12 days of clinical monitoring and intracranial stimulation.

### Clinical measures of migraine and associated comorbidities

We used the VAS score to evaluate moment-to-moment changes in attack severity. The 14-month preoperative headache diary recording and training of the recording paradigm in the patient’s first 2 days postoperatively ensured accuracy of the measurement of clinical symptoms. Microsoft Excel was used to collect and manage the study data. The patient underwent the entire procedure in our ward, remaining as calm and bedridden as possible to reduce disruption during the recording. During the clinical monitoring phase, considering that no further headache attacks had occurred, we induced migraine with the patient’s typical previous triggers. He recorded his VAS scores for 7 days in 5-minute increments and electrophysiological data in real time (**Figure 1b**). These data were classified into two types of symptoms, namely high and low symptoms. Furthermore, the aforementioned associated symptoms were recorded during headache attacks.

### Clinical-electrophysiological mapping via electrode stimulation

We tested the clinical effect of a set of stimulation parameters (10, 50, or 100 Hz; 300 µs; and 1–5 mA) through a systematic bipolar stimulation survey and blinded sham-controlled stimulation studies. The brain stimulation configuration was depicted by contact number and polarity (for example, 2-/3+ indicates that contact 2 is a cathode and contact 3 is an anode). The stimulation procedure was divided into two steps. First, we performed mapping of different parameters in all brain regions of interest to determine preliminarily the corresponding headache severity and other responses (**Figure 1c**). The process was repeated twice and guaranteed blind to the stimulation order, stimulation time, and other parameters. The candidate brain regions were chosen based on their ability to provide relief from headaches without eliciting severe discomfort responses. Second, we screened and validated the final stimulation targets from various perspectives, including different stimulation durations and blinding designs. The blinding method involved alternation of true and sham stimuli, disruption of the stimulus order, and other techniques.

### Signal processing

Real SEEG recordings were collected from the 14 SEEG electrodes with a sampling rate of 1000 Hz. All recordings were preprocessed with a 50-Hz comb notch filter to remove the line noise frequency and its harmonics, followed by a 0.5-to 300-Hz bandpass filter to eliminate baseline shifting; they were then re-referenced to the common average across all channels ^9^. To identify spectral biomarkers related to symptom severity states, 5-minute epochs after each self-reported VAS score were segmented for further spectral analyses. The spectral power for each channel was calculated at 30-second intervals and averaged over a 5-minute recording period using the Morlet wavelet transform (center frequency = 6 Hz). The seven frequency bands of interest were delta (δ, 1–4 Hz), theta (θ, 4–8 Hz), alpha (α, 8–12 Hz), beta (β, 12–30 Hz), low gamma (Lγ, 30–70 Hz), high gamma (Hγ, 70–150 Hz, and high-frequency oscillation (HFO) (150–300 Hz), yielding seven power features for each channel included (7 bands × 56 channels). Finally, the power feature for each brain region was determined by the spectral power averaged across all contacts within the brain regions (7 bands × 14 regions). All analyses were conducted using MATLAB R2021b (MathWorks, Natick, MA).

### Biomarker discovery for headache states representation

We determined the spectral power features of specific brain regions that were predictive of symptom severity using statistical analyses and cross-validated machine-learning models **(Figure 1d)**. First, to investigate the relationship between the derived spectral power features and VAS score, Spearman’s correlation coefficient (rho) along with its *p*-value were calculated for all features and montages. Furthermore, the two-sided Wilcoxon rank-sum test was used to validate whether a derived feature could significantly distinguish between low and high-symptom severity, yielding a *p*-value for each feature, which was considered significant at the α=0.05 level. Next, to minimize the high dimensionality of features and enhance the overall performance of the machine-learning classifier, the F-score feature selection method was implemented ^14^. The weight for a given feature was calculated using the F-score method. Greater weights contributed more to the migraine severity classification. Features with weights greater than 0.25 and *p*-values less than 0.05 were fed into different machine-learning classifiers, including decision trees (DT), the k-nearest neighbors (kNN) algorithm, logistic regression, Naive Bayes, and support vector machines (SVM). Their performances were assessed using the accuracy, sensitivity, specificity, F1-score, and area under the receiver operating characteristic (ROC) curve (AUC).

### Standard Protocol Approvals, Registrations, and Patient Consents

The Ethics Committee of the Chinese PLA General Hospital (S2022-384) approved the study. The patient provided written informed consent to participate in this research. The study was carried out in accordance with the World Medical Association’s Declaration of Helsinki.

## Data Availability

The datasets in the current study are available from the corresponding author on reasonable request. The source code used in this study is available from the corresponding author upon reasonable request.

## Results

### Outcomes after electrode implantation and removal

The patient reported complete pain relief after implantation of the SEEG electrodes, in contrast to the preoperative VAS scores of 4 to 7. When the clinical-electrophysiological recording was started, the patient used typical triggers according to the headache diary to provoke migraine attacks, with VAS scores of 1.5 to 7, which exhibited clinical features similar to those of past spontaneous migraine attacks. Following the clinical monitoring and mapping, the patient gradually experienced more spontaneous migraine attacks while waiting for the DBS procedure, as he had previously, but with a VAS score of only 1 to 2. He chose to postpone DBS implantation because of his satisfactory headache state. The preoperative dosage regimen was followed throughout the study and was still in use at the time of writing. During the 6-month postoperative follow-up, the patient’s headache attacks worsened in severity and frequency, becoming comparable to the preoperative headaches.

### High-frequency oscillation in the right nucleus accumbens as a crucial pain-related biomarker linked to pain fluctuations

The recordings were categorized into two groups (low and high migraine symptoms) according to the corresponding VAS score distribution (n = 53 and n = 45 events, respectively) (**Figure 2a**). Seven power characteristics were significantly correlated with and differed from the VAS score (**Table 1**), with HFO in the right nucleus accumbens (RNAc-HFO) being the most strongly correlated with the VAS score (rho = 0.5292, *p* < 0.0001) (**Figure 2b, c**) and significantly differing between the mild and severe pain levels (*p* = 0.0003) (**Figure 2d**). The characteristic ranking also revealed the greatest weighting of HFO in the RNAc brain region (**Figure 2e**). Features with corresponding weights of > 0.25 and both *p*1 and *p*2 values of < 0.05, as per the HFO of the RNAc, were selected as inputs to the classifier, which demonstrated that the linear support vector machine (SVM) outperformed the other classifiers, with accuracy and an area under the receiver operating curve remaining at 75.51% and 0.77, respectively (**Figure 2f**, **Table 2**). On the other side, power features from all brain regions were used as inputs to the machine-learning models. The results revealed that the linear SVM model was still optimal, with an accuracy of 71.43%; however, this accuracy was lower than that of the model using only the HFO-band power from the RNAc (**Table 3**). The correlation, significance analyses, and characteristic ranking of all contacts in the regions of interest supported the optimal discrimination of HFOs on the four contacts of the RNAc (**Figure 2g**, **Table 4**).

**Figure 2.**
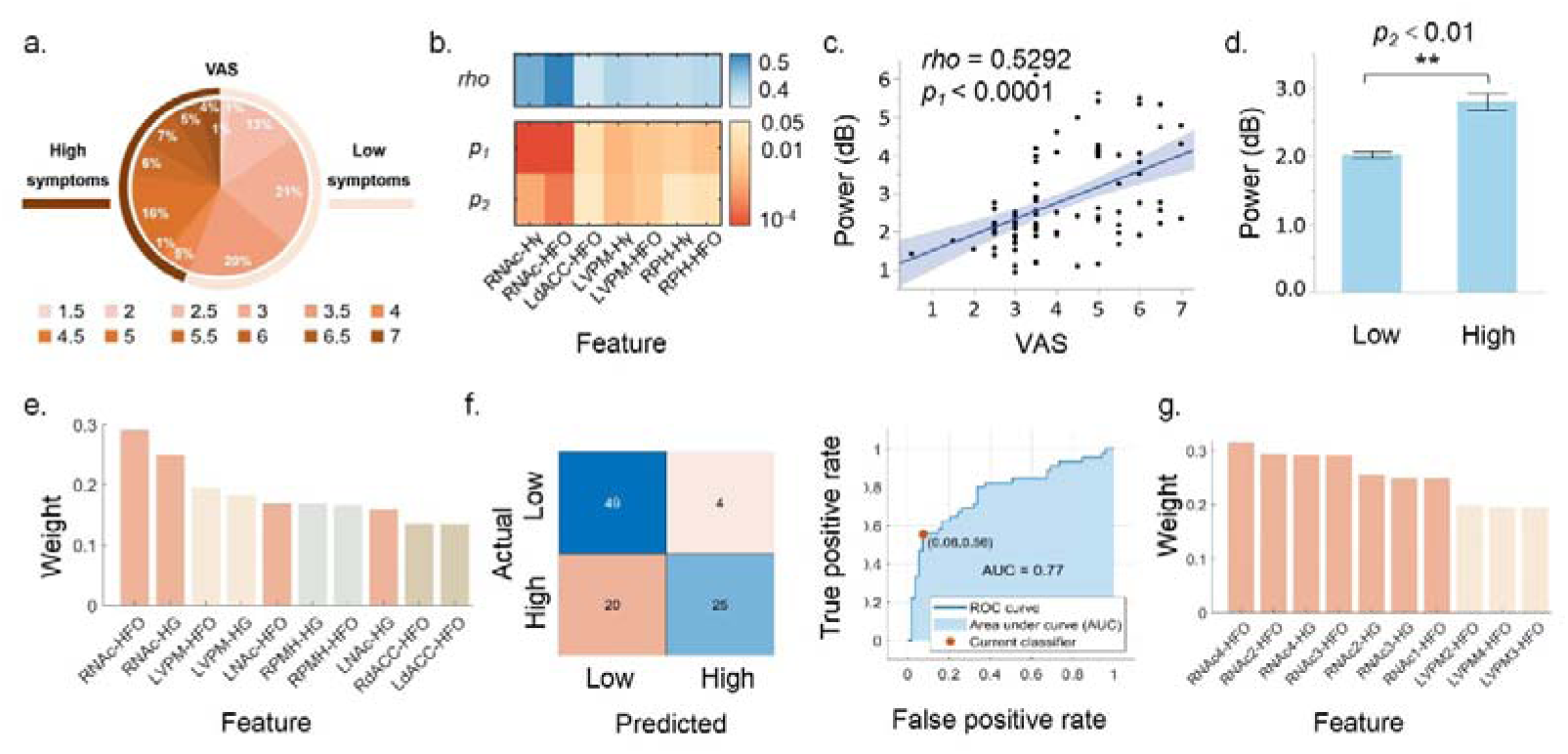
**Identification of pain-related biomarkers. a**. VAS scores during the monitoring ranged from 1.5 and 7. Data were divided into high and low-symptom groups according to a VAS cut-off score of 3.5 (*n* = 53 and *n* = 45 events, respectively). **b**. The relationship and discrimination of spectral power features and VAS scores. RNAc-HFO had the most striking features among the seven power characteristics that were significantly associated with and distinct from the VAS. rho, *p*_1_: Spearman’s correlation coefficient along with its p-value between power and VAS scores; *p*_2_: Wilcoxon rank-sum test result representing the difference in power between the low and high migraine symptom groups. **c**. Correlation of RNAc-HFO band power and VAS scores. Each point represents a VAS score and the corresponding RNAc-HFO band power for a specific trial (total of 98 data points). **d**. The ability of the RNAc-HFO band power to discriminate between high and low-symptom groups. Bar heights indicate mean values and black error bars indicate the standard errors. *\*p* < 0.05, \*\**p* < 0.01, \*\*\**p* < 0.001. **e**. Ranking of all extracted features using the F-score feature selection algorithm, with the highest weight indicating the greatest ability to discriminate between the high and low-symptom groups. The greatest headache discriminatory ability was demonstrated by RNAc-HFO. HG: High gamma. **f**. Cross-validated machine-learning models. Features with weights > 0.25 and *p*_1_ and *p*_2_ < 0.05 were chosen as input to the classifier (the HFO band power in the RNAc), with the confusion matrix and ROC curve representing the performance of the optimal classifier (SVM). **g**. Ranking of contact features using the F-score feature selection algorithm, and the HFO of the various contacts of the RNAC all had higher weights.

**Table 1.** Significant power characteristics statistics in different brain regions

|  | RNAC | RNAC | LdACC | LVPM | LVPM | RPH | RPH |
| --- | --- | --- | --- | --- | --- | --- | --- |
|  | Hγ | HFO | HFO | Hγ | HFO | Hγ | HFO |
| *rho* | 0.4786 | 0.5292 | 0.3721 | 0.4198 | 0.4070 | 0.3993 | 0.4114 |
| *p<sub>1</sub>* | <0.0001 | <0.0001 | 0.0181 | 0.0019 | 0.0036 | 0.0052 | 0.0029 |
| *p<sub>2</sub>* | 0.0012 | 0.0003 | 0.0408 | 0.0024 | 0.0044 | 0.0376 | 0.0187 |

\* *rho*, *p<sub>1</sub>*: Spearman correlation, representing correlation between power and VAS scores; *p<sub>2</sub>*:
Wilcoxon rank sum test, representing variability of power between low and high migraine symptoms. Hγ: High gamma; HFO: High-frequency oscillation.

**Table 2.** Comparisons of the performances of different classifiers using features of all brain regions

| Classifier | Acc. | Spec. | Sen. | F1- score | AUC |
| --- | --- | --- | --- | --- | --- |
| DT | 67.35% | 71.70% | 62.22% | 63.64% | 0.70 |
| LDA | 74.49% | 90.57% | 55.56% | 66.67% | 0.77 |
| LR | 72.45% | 86.79% | 55.56% | 64.94% | 0.77 |
| NB | 74.49% | 90.57% | 55.56% | 66.67% | 0.76 |
| SVM | **75.51%** | 92.45% | 55.56% | 67.57% | 0.77 |

\*Features with weights greater than 0.25, and both p1 and p2 values less than 0.05 were selected as input to the classifiers (the HFO band power in the RNAc). Linear SVM outperformed other classifiers when using only the HFO band power of the RNAc for classification, with accuracy, specificity, sensitivity, F1-score, and AUC remaining at 75.51%, 92.45%, 55.56%, 67.57%, and 0.77, respectively, followed by linear discriminant and plain Bayes with 74.49% accuracy.

**Table 3.** Comparisons of the performances of different classifiers using features of different contacts in brain regions

| Classifier | Acc. | Spec. | Sen. | F1- score | AUC |
| --- | --- | --- | --- | --- | --- |
| DT | 63.39% | 75.47% | 62.22% | 65.12% | 0.68 |
| LDA | 67.35% | 64.15% | 71.11% | 66.67% | 0.68 |
| LR | 56.12% | 50.94% | 62.22% | 56.57% | 0.58 |
| NB | 59.18% | 50.94% | 68.89% | 60.78% | 0.77 |
| SVM | **71.43%** | 77.36% | 64.44% | 67.44% | 0.76 |

\*Features of different contacts in brain regions were selected as input to the classifiers, and linear SVM outperformed other classifiers with accuracy, specificity, sensitivity, F1-score, and AUC remaining at 71.43%, 77.36%, 64.44%, 67.44%, and 0.76, respectively.

**Table 4.** Significant power characteristics statistics in different contacts in brain regions

|  | RNAc-1 | RNAc-2 | RNAc-2 | RNAc-3 | RNAc-3 | RNAc-4 | RNAc-4 |
| --- | --- | --- | --- | --- | --- | --- | --- |
| | HFO | Hγ | HFO | Hγ | HFO | Hγ | HFO |
| *rho* | 0.4511 | 0.4947 | 0.5367 | 0.4953 | 0.5300 | 0.5289 | 0.5449 |
| *p*<sub>1</sub> | 0.0011 | 0.0001 | < 0.0001 | < 0.0001 | < 0.0001 | < 0.0001 | < 0.0001 |
| *p*<sub>2</sub> | 0.0145 | 0.0033 | 0.0008 | 0.0041 | 0.0015 | 0.0004 | 0.0005 |
|  | LVPM-1 | LVPM-2 | LVPM-2 | LVPM-3 | LVPM-3 | LVPM-4 | LVPM-4 |
| | Hγ | Hγ | HFO | Hγ | HFO | Hγ | HFO |
| *rho* | 0.4143 | 0.4002 | 0.3954 | 0.4149 | 0.4108 | 0.4294 | 0.4334 |
| *p*<sub>1</sub> | 0.0010 | 0.0199 | 0.0250 | 0.0097 | 0.0119 | 0.0046 | 0.0037 |
| *p*<sub>2</sub> | 0.0276 | 0.0164 | 0.0223 | 0.0113 | 0.0164 | 0.0044 | 0.0085 |

\* Hγ : High gamma; HFO: High-frequency oscillation.

### Left dorsal anterior cingulate cortex as an optimal target for personalized DBS

Different stimulus parameters elicited varying responses in different regions, including pain responses and other associated symptoms; these included typical accompanying symptoms, autonomic symptoms, and olfactory hallucination, all of which were associated with the patient’s previous migraine attacks. Clinically, the patient reported a reduction in headache severity during stimulation in two brain regions (left dorsal anterior cingulate cortex [LdACC] and RNAc), with varying degrees of headache exacerbation in most other brain regions (**Figure 3a**). Based on the VAS scores reported during the 5-day stimulation period, we identified the LdACC as the most effective stimulation target (**Figure 3b**). Pre-and post-stimulus VAS scores showed significantly greater improvement (*p* = 0.006) when compared with the RNAc (*p* = 0.081) (**Figure 3c**). In terms of electrophysiology, RNAc-HFO was consistently lower after than before stimulation in all pain relief trials, validating the effectiveness of HFO in the RNAc as a biomarker of pain severity (**Figure 3d**). Among these effects, the stimulation of LdACC produced a significant difference in electrophysiological effects (*p* < 0.01) (**Figure 3e**). Furthermore, the low-frequency phase of the LdACC had a significant modulatory effect on the HFO band amplitude of the RNAc, which was increased in the high-symptom state. This confirmed the modulatory effect of the LdACC on the RNAc during headache fluctuation (**Figure 3f**).

**Figure 3.**
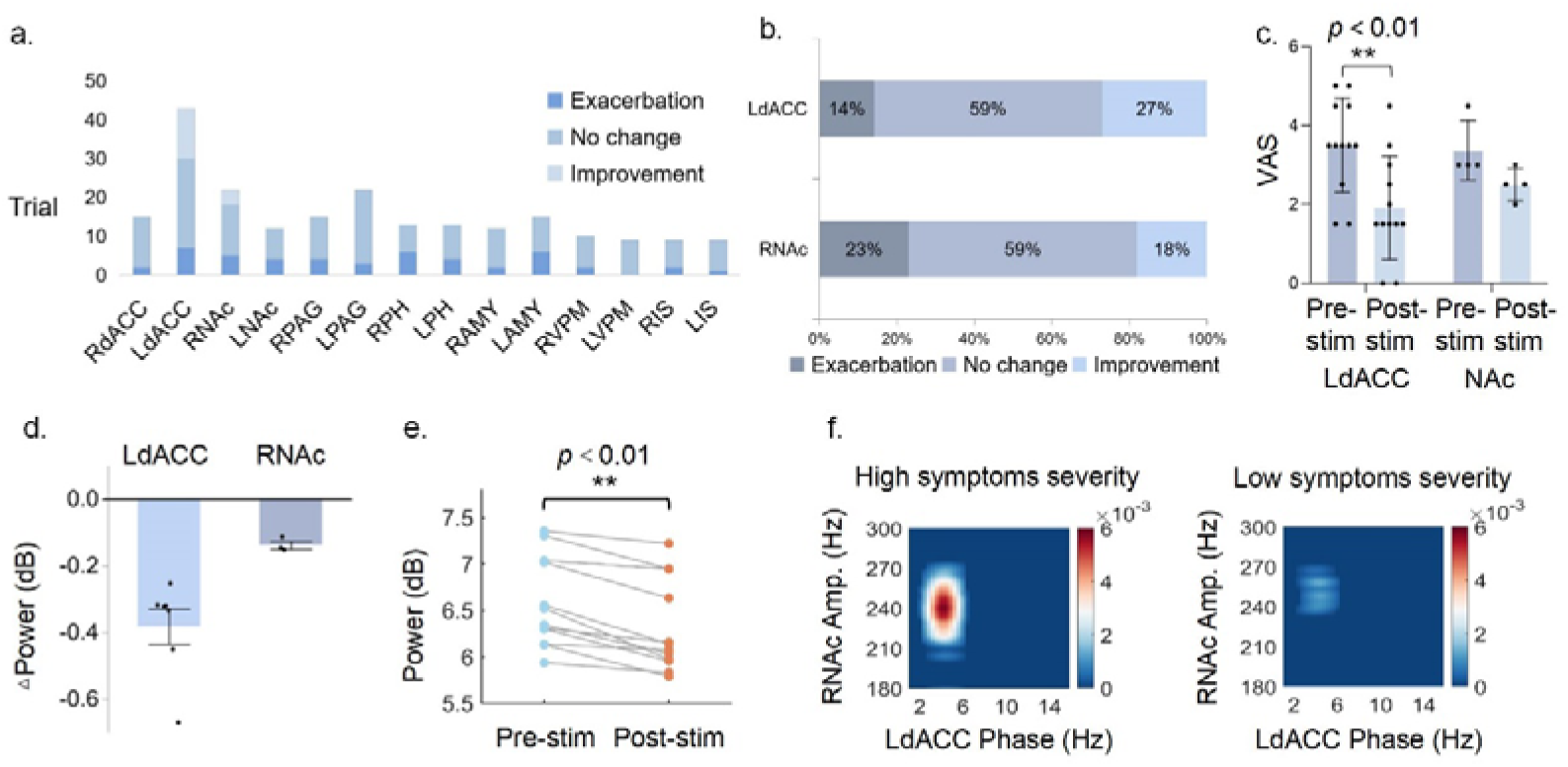
**Identification of the optimal stimulation target. a**. Pain responses to various stimuli in various regions. Pink represents aggravation, while blue represents relief. Only two brain regions, RNAc and LdACC, were stimulated, and headache intensity improved. **b**. Stimulation responses of the two optional regions, i.e., improvement, unchanged, or exacerbation. **c**. Distribution of VAS scores pre-and post-stimulation in LdACC and RNAc, with a statistically significant difference in the former. \*\**p* < 0.01. **d**. Validation of biomarker against data with headache improvement in response to stimulation, with greater pre-and post-stimulus in LdACC. **e**. Statistical analysis of RNAc-HFO power variation pre-and post-stimulation in LdACC before and after stimulation on data with headache improvement. In data where stimulation caused headache improvement, there was a decrease in the corresponding biomarker power. \*\**p* < 0.01. **f**. Verification of functional modulation between RNAc and LdACC using phase-amplitude coupling (PAC). The low-frequency phase of LdACC had a great modulatory effect on the HFO band amplitude of RNAc, which was enhanced in the high-symptom state.

## Discussion

In this study, we developed a personalized strategy through multi-dimensional and multi-perspective analysis. First, we tracked the baseline period of the patient’s headache attacks using VAS scores and electrophysiology, to characterize the various headache states. Based on the clinical state of “low” or “high” symptoms, the corresponding electrophysiology data were combined to identify the biomarker that characterized the optimal state for this patient, which was originally explored in previous migraine and EEG studies ^15, 16^. On the one hand, biomarker research facilitates the identification of personalized key targets against the complexity of individual brain networks due to pathophysiology. On the other hand, the current trend in DBS implementation is the initiation of biomarker-based on-demand stimulation via biomarker monitoring ^5^. Second, in terms of therapeutic targeting, we first intuitively identified the best stimulus target for patient comfort using personalized stimulus-response mapping. When improvement was noted in the subjective headache symptom ratings, which is an important indicator of efficacy, we could identify the target of stimulation to some extent. However, because subjective feelings have certain confounding factors, such as emotions and perceptions, we further validated this with objective data drivers. In conjunction with the biomarker identified in the first step, we paired the monitoring state signal with clinical symptoms and successfully validated the relationship between a decline in the biomarker and headache improvement, to objectively determine the physiological effect of the stimulus target. In addition, phase-amplitude coupling in this study was used to validate the functional modulation between the biomarker and stimulated regions, confirming that the modulation between the two is an important link in clinical effects. Although the final stimulation efficacy is subject to further investigation, the multidimensionally validated trial protocol ensures the best possible treatment.

In the present study, RNAc-HFO optimally reflected pain fluctuations with a positive association. Previous research has suggested that the NAc could be used as a biomarker for pain or migraine, with a decrease in the volume of the RNAc as shown through neuroimaging studies ^17^ and increased functional connectivity in the RNAc with some brain regions in patients with migraine ^18^. Iron deposition in NAC may be a biomarker for migraine chronicity and migraine-related dysfunctions^19^. Furthermore, we identified the LdACC as the most effective target region for the patient based on his clinical response and further validation of biomarkers that could be improved, which is an important step in personalized treatment. Some studies have shown that functional changes in the ACC (anterior cingulate cortex) might serve as a biomarker for migraine prevention. Changes in cortical thickness in the left posterior cingulate ^20^, increases in gamma-aminobutyric acid levels in the ACC or posterior cingulate ^21, 22^, and the blood oxygenation level-dependent response in the perigenual region of the right ACC ^23^ have all been linked to clinical aspects of migraine such as headache frequency and intensity. These findings support our hypothesis that the ACC might be used as a potential therapeutic target for migraine. Furthermore, the modulatory effects of electrical stimulation in the ACC and RNAc-HFO on migraine in our study demonstrated that functional regulation between the ACC and NAc may be involved in migraine pathophysiology. Optogenetics have revealed that the ACC projection to the NAc pathway is selectively involved in pain and analgesia social transfer, implying that the NAc is a downstream target of ACC pain modulation ^24^. The ACC most likely mediates pain-related aversive behavior by projecting to NAc D2-type medium spiny neurons and other mesolimbic dopamine systems ^25^. Thus, we hypothesize that a similar pathway exists in migraine, which supports the feasibility of the LdACC as a therapeutic target in our study. However, specific molecular mechanisms should be investigated further. More importantly, the effectiveness must be validated based on the patient’s follow-up outcome after DBS placement.

Interestingly, the patient reported an approximately 2-week absence of migraine attack after SEEG implantation; the migraine attacks then gradually returned during the subsequent follow-up. Given the results of previous studies on SEEG or DBS implantation, we speculate that several factors may be at work in this phenomenon. First, the microlesion effect (MLE), which typically lasts 2 to 4 weeks, may result in a reduction or even disappearance of clinical symptoms; this is primarily due to reversible minor tissue exudation and edema around the target region ^26^. The MLE has been shown to be a predictor of DBS efficacy and to aid in the indirect assessment of whether the electrodes are in the proper target position ^27^. Thus, the MLE at the LdACC may have temporarily relieved our patient’s migraine, corroborating the validity of the subsequent stimulation mapping. Second, some research has suggested that subanesthetic doses of propofol could be used in the acute or prophylactic treatment of intractable migraine, with efficacy lasting up to 6 months ^28, 29^; however, this needs to be confirmed. The intraoperative maintenance of anesthesia in our patient had the potential to result in headache relief followed by recurrence of migraine 2 weeks later. However, the validity of this conjecture is difficult to confirm based on the available information. Furthermore, the placebo effect is important in both DBS ^26, 30^ and SEEG implantation. In fact, some studies have shown that more invasive procedures have a larger placebo effect ^31^; thus, we cannot completely rule out the possibility of a placebo effect causing bias in the patient’s assessment of symptoms. However, the placebo-controlled blind stimulation procedures as well as the double-blind stimulation recording and data analyses in our study are expected to have reduced the placebo effect in our patient.

This is the first study to explore personalized targets for DBS treatment of rCM to date. However, it has several limitations. First, because this was an individualized design for one patient, the results cannot be generalized to other patients with migraine. Second, the calibration of the clinical recording and stimulation responses may be influenced by certain subjective factors, such as emotions; therefore, we trained the patient in advance for recording and increased the number of trials to ensure accuracy. Third, we used different stimulus durations in our experimental design to investigate the patient’s recordings of his headaches, but the optimal stimulation parameters for headache response in practice require further investigation. Furthermore, some therapeutic regimens, such as calcitonin gene-related peptide (CGRP antagonists and monoclonal antibodies to CGRP or its receptor) were not available in China before the study and could not be administered to our patient; as a result, their efficacy remains unknown. Finally, because the patient was highly satisfied with his clinical condition at the time of SEEG electrode removal, the second step of DBS implantation was postponed and will be scheduled later based on his condition and personal preferences.

## Conclusions

This is the first study to evaluate SEEG recordings in a patient with migraine, to deliver precise and personalized DBS. To provide specialized care, we identified the LdACC as a therapeutic target for DBS using the biomarker of HFO in the RNAc. To determine the efficacy of this treatment approach, we plan to perform DBS implantation and monitor this patient for a long time. Our preliminary study conclusively shows that this unique concept for tailored DBS in patients with rCM is feasible, and we anticipate further verification of its precise therapeutic use in various forms of headaches and even pain.

## Competing interests

The authors declare the existence of a non-financial competing interest.

## Data Availability

All data produced in the present study are available upon reasonable request to the authors.

## Acknowledgment

Thanks to the patient’s cooperation in this study.

## Funding

This work was supported by the National Natural Science Foundation of China (Grants 82171208) and Health Special Research Projects (Grants 22BJZ21).

## Notes

### Competing Interest Statement

The authors have declared no competing interest.

### Clinical Trial

ChiCTR2200062135

### Author Declarations

The Ethics Committee of the Chinese PLA General Hospital (S2022-384) approved the study.

### Summary of Updates

Author affiliations and E-mail updated.

